# Vitamin D status in chronic tic-disorder and comorbid obsessive-compulsive disorder and attention-deficit/hyperactivity disorder: A pan-European study

**DOI:** 10.1101/19012062

**Authors:** Molly Bond, Natalie Moll, Alicia Rosello, Rod Bond, Jaana Schnell, Bianka Burger, Pieter J. Hoekstra, Andrea Dietrich, Anette Schrag, Eva Kocovska, Davide Martino, Norbert Mueller, Markus Schwarz, Ute-Christiane Meier, the EMTICS collaborative group

## Abstract

**Background:** Hypovitaminosis-D has been linked to neuropsychiatric conditions, but little is known about vitamin D status in individuals with chronic tic disorders (CTD). This study sought to determine whether vitamin D levels are associated with the severity of tics and presence and severity of comorbid obsessive-compulsive disorder (OCD) and/ or attention-deficit/hyperactivity disorder (ADHD).

**Methods:** This is a cross-sectional sub-study of the European Multicentre Tics in Children Studies (EMTICS). Vitamin D [25(OH)D] levels (ng/ml) were obtained for 327 participants with CTD at study entry and tic severity was measured using the Yale Global Tic Severity Scale (YGTSS). An association between tic severity and presence and severity of comorbid OCD and/or ADHD symptoms was analysed using multilevel models controlling for season, site, sex, age and presence of comorbid OCD and/or ADHD.

**Results:** The participants comprised 247 boys and 80 girls (4-16 years, mean [SD], 10.9 [2.72]). Hypovitaminosis-D ([25(OH)D] < 20ng/ml) was present in 33% of participants with CTD and/ or comorbid OCD and/ or ADHD. The authors found no association between 25(OH)D levels and tic severity or OCD symptom severity. However, lower 25(OH)D levels were significantly associated with increased severity of ADHD symptoms in participants with CTD (*β* = -0.25, s.e. = 0.08, 95% CI, -0.42 to -0.09; *p* < 0.01).

**Conclusion:** Additional research is needed to confirm these findings and assess whether hypovitaminosis-D may represent an underlying biological vulnerability for comorbid ADHD symptoms in CTD as it may signpost novel strategies for prevention and treatment.

## INTRODUCTION

Chronic tic disorders (CTD) are neurobehavioral disorders with an onset in childhood or adolescence. Tourette Syndrome (TS), perhaps the most widely known CTD, is characterised by the combination of multiple motor tics and at least one vocal tic that persist for more than a year ^1^ and affects around 0.8% of children worldwide ^2 3^. A large proportion of individuals with CTD have associated neuropsychiatric comorbidities such as obsessive-compulsive disorder (OCD) and attention-deficit/hyperactivity disorder (ADHD) ^3^.

Genetic factors are important in determining susceptibility to TS, OCD and ADHD as highlighted by a concordance rate for monozygotic twins with TS of 53% ^3-5^, OCD of 50% ^6^ and ADHD of 70-80% ^7^. However, the fact that monozygotic twins often share an environment and estimates of heritability are less than 100% in all these conditions asserts quite strongly that environmental factors are also involved. Their identification is, therefore, of great interest as they may be modifiable in contrast to genetic susceptibility and signpost novel strategies for prevention and treatment.

One environmental factor of interest is vitamin D, a neurosteroid hormone with pleiotropic effects, which plays an important role in skeletal health but also in neural and immune functioning ^8 9 10^. Vitamin D is a fat-soluble vitamin that is naturally present in few foods and mainly produced in skin during exposure to sunlight (UVB radiation). Its synthesis is therefore greatly influenced by season, latitude, air pollution, skin pigmentation, sunscreen use and aging, as well as by genetic factors, altered absorption/metabolism and medication ^11 9^. Vitamin D influences a large number of biological pathways, which may help explain association studies relating hypovitaminosis-D with increased risk for many chronic diseases ^11 8 9^.

A number of studies have drawn a link between hypovitaminosis-D and autoimmune diseases (e.g. multiple sclerosis, Type 1 diabetes mellitus), metabolic disorders (e.g. atherosclerosis) and neoplastic disorders (e.g. colon cancer) ^12 11^. Hypovitaminosis-D has more recently also been associated with several neuropsychiatric diseases such as mood disorders, schizophrenia, autism, Parkinson’s disease and Alzheimer’s disease ^8 9 10^.

To date, epidemiological evidence on the association between vitamin D and tic disorders is limited. One study assessed vitamin D status in Chinese children (n = 179) with tic disorder without psychiatric comorbidities and reported hypovitaminosis-D and an inverse correlation between vitamin D levels and tic severity (r = -0.32) ^13^. We, therefore, wanted to test for such an association in our large population sample of children and adolescents with CTD and comorbid OCD and/or ADHD, which was collected as part of the European Multicentre Tics in Children Studies (EMTICS).

Our hypothesis is that hypovitaminosis-D is associated with the severity of clinical features and psychiatric comorbidities in children and adolescents with CTD. To our knowledge this is the first study investigating the relationship between vitamin D (25(OH)D) status and the severity of CTD and presence and severity of comorbid OCD and/or ADHD in a pan-European cohort ^14^.

## MATERIALS AND METHODS

### Participants

The participants were part of EMTICS (n = 715), a prospective European longitudinal observational study involving 16 centres located in nine countries across Europe and in Israel ^14^. Our cross-sectional study measured 25(OH)D levels at study entry of 327 children and adolescents (aged 4 to 16 years) with TS or another CTD according to DSM-IV-TR (APA, 2000). The study cohort represented a selected sub-sample of the EMTICS cohort based on availability of serum samples. The study was approved by the Ethics Committees at all participating clinical centres and samples and data were obtained with informed consent by the parents or legal guardians and written or oral assent by the participating child before entering the study.

### Clinical assessment

An established diagnosis of TS, chronic motor or chronic vocal tic disorder was confirmed by study clinicians according to DSM-IV-TR criteria ^15^ using semi-structured interviews (Table 1b). Lifetime presence of OCD or ADHD was also assessed according to DSM-IV-TR criteria ^15^.

**Table 1:**
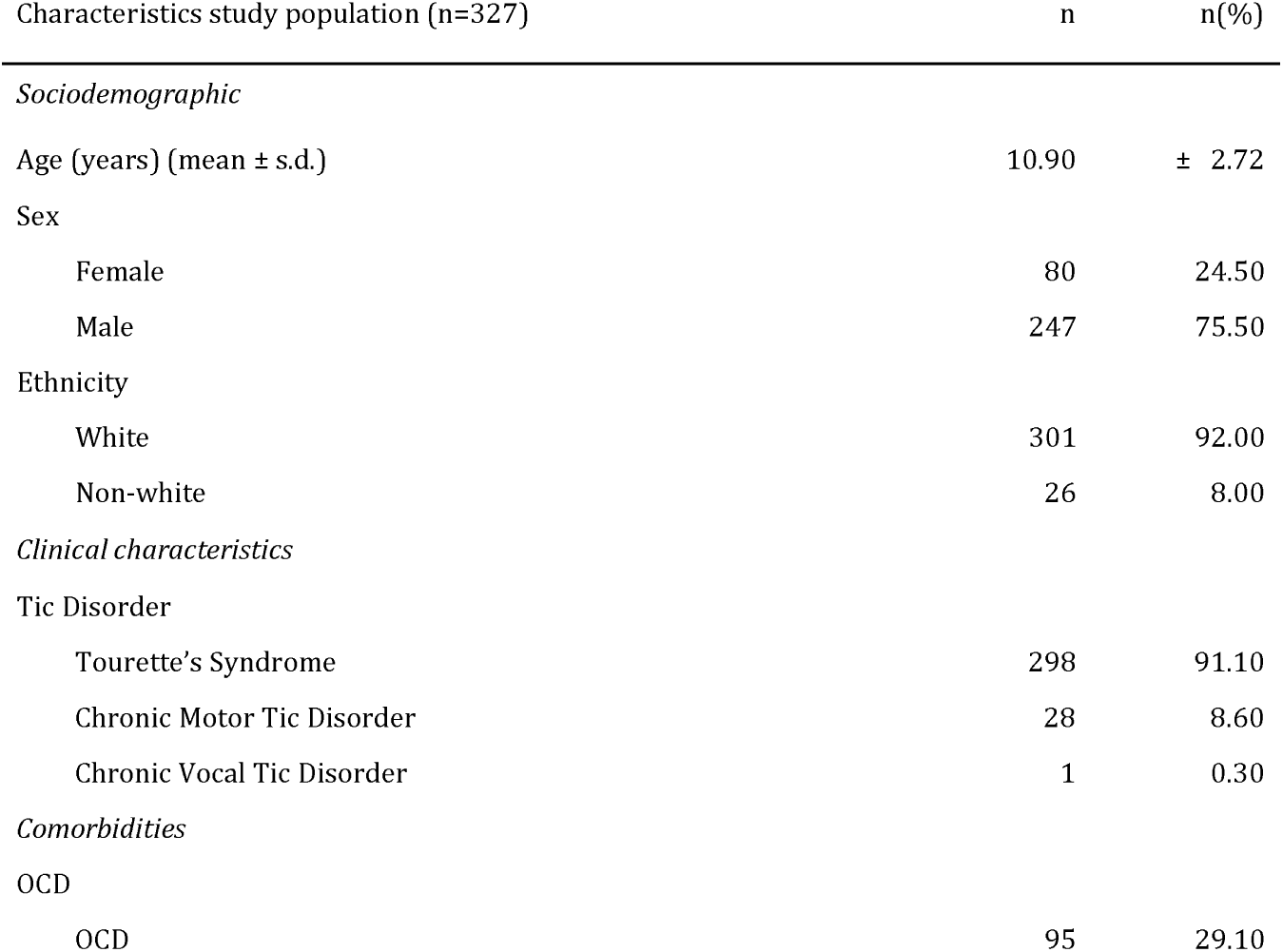
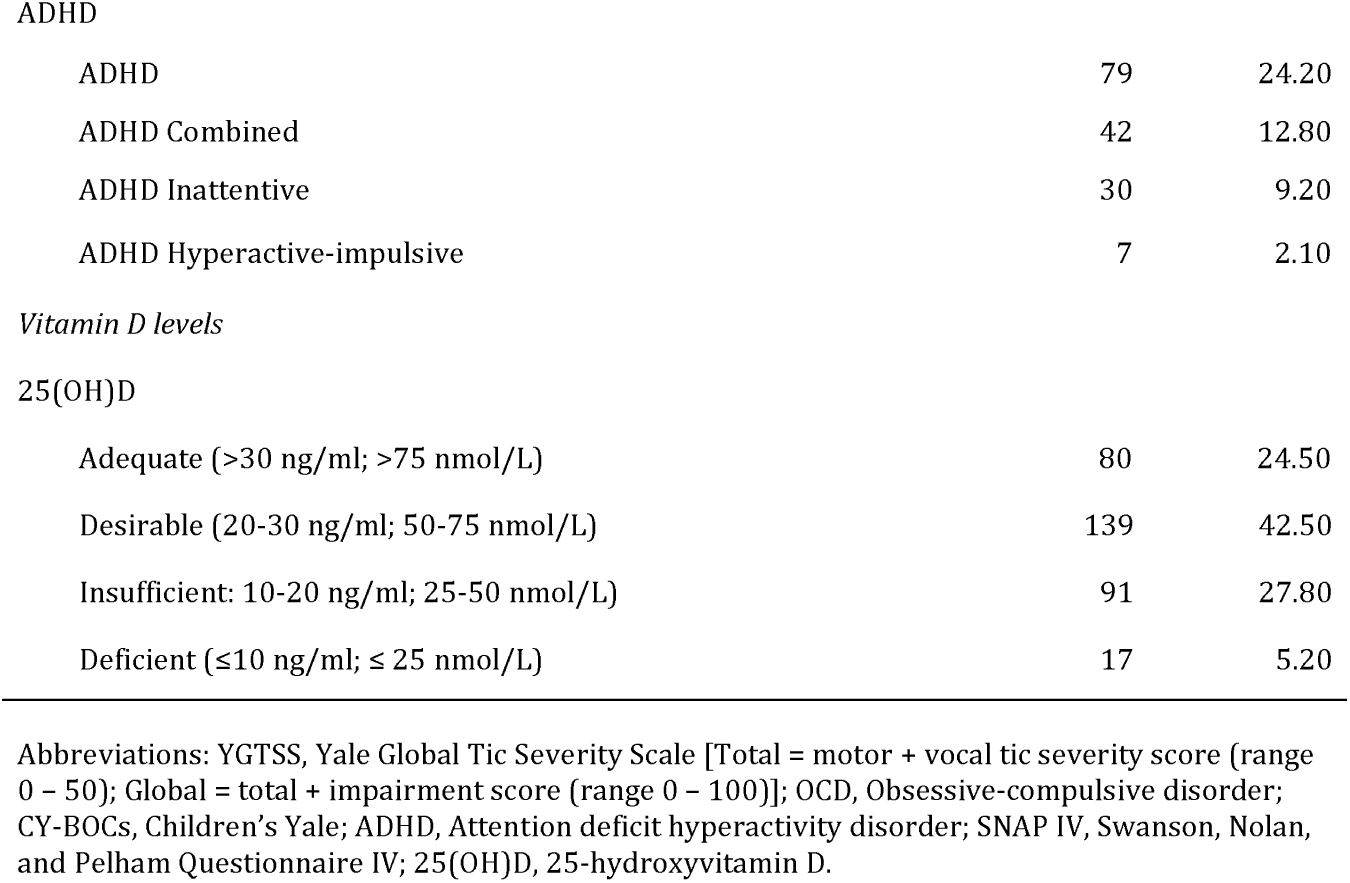
Demographic Statistics

| Characteristics study population (n=327) | n | n(%) |
| --- | --- | --- |
| *Sociodemographic* |  |  |
| Age (years) (mean ± s.d.) | 10.90 | $\pm 2.72$ |
| Sex |  |  |
| Female | 80 | 24.50 |
| Male | 247 | 75.50 |
| Ethnicity |  |  |
| White | 301 | 92.00 |
| Non-white | 26 | 8.00 |
| *Clinical characteristics* |  |  |
| Tic Disorder |  |  |
| Tourette's Syndrome | 298 | 91.10 |
| Chronic Motor Tic Disorder | 28 | 8.60 |
| Chronic Vocal Tic Disorder | 1 | 0.30 |
| *Comorbidities* |  |  |
| OCD |  |  |
| OCD | 95 | 29.10 |
| ADHD |  |  |
| ADHD | 79 | 24.20 |
| ADHD Combined | 42 | 12.80 |
| ADHD Inattentive | 30 | 9.20 |
| ADHD Hyperactive-impulsive | 7 | 2.10 |
| 25(OH)D |  |  |
| Adequate (>30 ng/ml; >75 nmol/L) | 80 | 24.50 |
| Desirable (20-30 ng/ml; 50-75 nmol/L) | 139 | 42.50 |
| Insufficient: 10-20 ng/ml; 25-50 nmol/L) | 91 | 27.80 |
| Deficient ($\leq 10$ ng/ml; $\leq 25$ nmol/L) | 17 | 5.20 |

Abbreviations: YGTSS, Yale Global Tic Severity Scale [Total = motor + vocal tic severity score (range 0 – 50); Global = total + impairment score (range 0 – 100)]; OCD, Obsessive-compulsive disorder; CY-BOCs, Children's Yale; ADHD, Attention deficit hyperactivity disorder; SNAP IV, Swanson, Nolan, and Pelham Questionnaire IV; 25(OH)D, 25-hydroxyvitamin D.

Our main outcome measure for tic severity was the Yale Global Tic Severity Scale (YGTSS), the gold standard to rate tic severity ^16^. The YGTSS was assessed by study clinicians and comprised the YGTSS Total (motor + vocal tic severity; range 0 – 50) and YGTSS Global score (total + impairment score; range 0 – 100). Our primary outcome for analysis was YGTSS Total score. In addition, the well-validated Clinical Global Impression Scale Severity (CGI) was also used to assess tic severity during the previous week ^17^.

The Children’s Yale Brown Obsessive–Compulsive Scale (CY-BOCS) was used to rate obsessive-compulsive symptom severity. The CY-BOCS total score (range 0-40) is comprised two subscales: obsessions (range 0-20) and compulsions (range 0-20) ^18^. The parent-reported Swanson, Nolan and Pelham-version IV rating scale (SNAP-IV) assessed ADHD symptom severity ^19^. We used subscale severity scores to assess differences between inattentive (range 0-27) and hyperactivity-impulsive symptom severity (range 0-27). We also used ADHD symptom count based on the DSM-IV-TR (range 0-18) and assessed subscales for inattentive symptoms (range 0-9) and hyperactivity/ impulsivity (range 0-9).

### Analysis of 25-hydroxyvitamin D (25(OH)D)

Serum samples of 327 participants with CTD were collected by the clinical centres and sent to the University Hospital Munich (LMU). The samples were stored at -80°C until analysis. The level of 25-hydroxyvitamin D [25(OH)D] was measured in the ISO 15189 accredited lab on a DiaSorin Liaison analyser by chemiluminescent immunoassay technology. Serum samples were processed according to the manufacturer’s instructions. The range of the assay was 4 ng/mL (10 nmol/L) up to 150 ng/mL (375 nmol/L).

In our study, hypovitaminosis-D was reported at levels under 20 ng/ml (50 nmol/L) in accordance with the US Endocrine Society ^20^. The Institute of Medicine has set the optimal serum 25(OH)D level at ≥20 ng/ml (≥50 nmol/L) ^21^. A serum 25(OH)D lower than ≤10 ng/ml (≤25 nmol/L) is considered deficient, as it may be associated with skeletal disease, and levels from 10-20 ng/ml (25-50 nmol/L) are considered insufficient. In accordance with some experts levels of 25(OH)D >30 ng/ml (>75 nmol/L) are considered as optimal ^20^.

### Statistical Analysis

We used multilevel models to investigate the association of 25(OH)D levels and tic severity (YGTSS scores and CGI score) and severity of comorbid OCD (CY-BOCS scores) and ADHD (SNAP-IV and DSM-IV-TR) symptoms. OCD and ADHD symptom severity scores were taken from the entire cohort and not just those with a comorbid diagnosis. For binary outcome variables, we used generalised linear mixed models to assess whether 25(OH)D levels influenced the probability of an OCD or ADHD diagnosis. Since this is a multicentre study, the data are hierarchically structured whereby participants are clustered within site. We therefore used multilevel models with site as the cluster variable in these analyses to control for any between site differences, which includes latitude of centre.

To correct for seasonal variation in 25(OH)D levels, we used a previously described method ^22^. In short, it involves a sinusoidal regression model whereby the following equation predicts 25(OH)D levels for any given day of the year (*y* = 25(OH)D level and *t* = day of year e.g. for January 1^st^, *t* = 1):

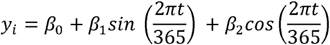

The difference between predicted 25(OH)D on a given day and the individual’s actual level gives a deseasonalised residual value of 25(OH)D controlling for season. We included each participants’ serum 25(OH)D concentration adjusted for seasonal variation, as well as the average adjusted serum 25(OH)D by site as predictors of tic severity and comorbidity severity in order to disentangle the contextual effects from the person-level effects ^23 24^. Sex, age and the presence of a comorbidity (OCD and/ or ADHD) were also entered as covariables.

To estimate the prevalence of hypovitaminosis-D throughout the year, we added the predicted value of when 25(OH)D levels would be highest and at the mid-point in the year to each participants’ residual 25(OH)D value from the sinusoidal regression model for seasonal variation to estimate the distribution of 25(OH)D at these points in the year. This gave an estimation of the proportion of participants who would present with hypovitaminosis-D throughout the year, as well as for six months of the year.

## RESULTS

### Study cohort

The study included 247 males (75.5%) and 80 females (24.5%) aged between 4.54 and 16.99 (mean [SD], 10.90 ± 2.72) (Table 1, Figure 1A). The majority of participants were of European white Caucasian ancestry (n = 301, 92%), whilst the remaining (n = 26, 8%) were mixed, Middle Eastern, North African, Asian or of unknown ancestry. Upon enrolment into the study, most participants were diagnosed with TS (n = 298; 91.1%) and a minority with a motor CTD (n = 28) or vocal CTD (n = 1).

**Figure 1:**
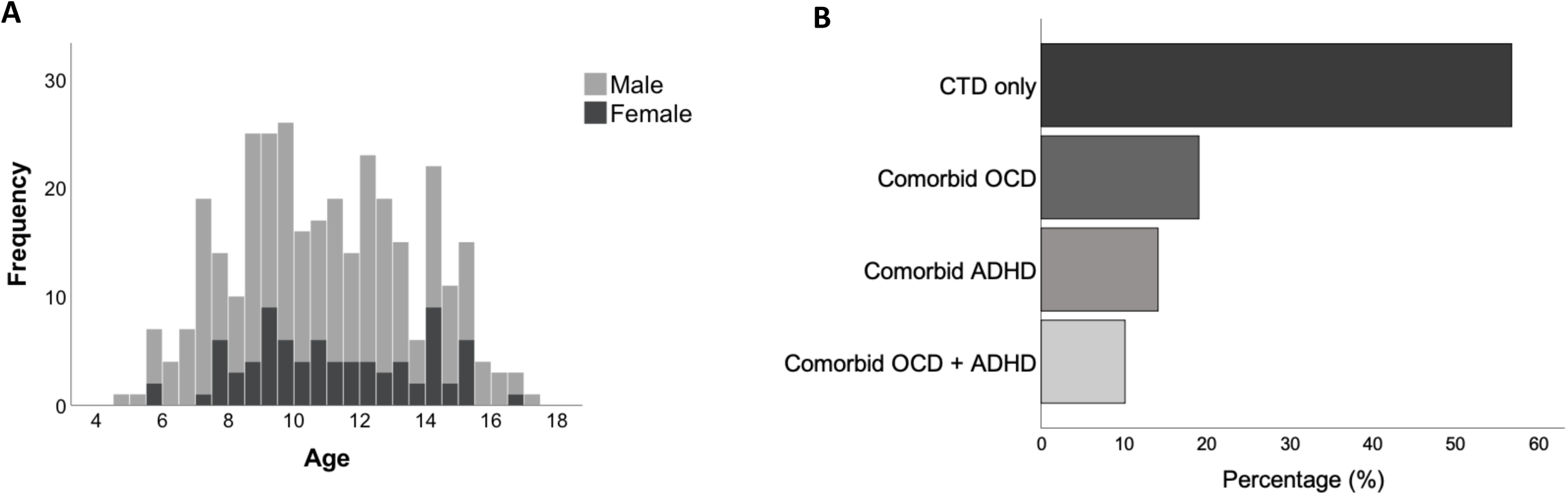
(A) Distribution of gender and age of young individuals with chronic tic disorder (CTD). The graph shows the frequency of male (grey) and female (black) CTD patients according to age. **(B) Prevalence of comorbidities of young individuals with chronic tic disorder (CTD)**. The graphs show the percentage of study participants with CTD (56.6%) and comorbid obsessive compulsive-disorder (OCD; 19%) and/or attention-deficit hyperactivity disorder (ADHD; 14.1%) and comorbid OCD and ADHD (10.1%).

### Prevalence of comorbid disorders

The prevalence of comorbidities was high (n = 141, 43.3%) (Table 1, Figure 1B). The most prominent comorbidity was OCD (n=95; 29.0%) followed by ADHD (n=79; 24.0%) (Table 1) and, of these, 10.1% presented with both comorbid OCD and ADHD (Figure 1B).

### Hypovitaminosis-D

Vitamin D status was assessed by measuring circulating serum levels of 25(OH)D, which is a summation of cutaneous synthesis from solar exposure and dietary sources ^25^. Levels of 25(OH)D fluctuated within the cohort according to season following the sine curve used to correct for seasonality (Figure 2A). Highest levels were found during late summer and lowest levels in early spring.

**Figure 2:**
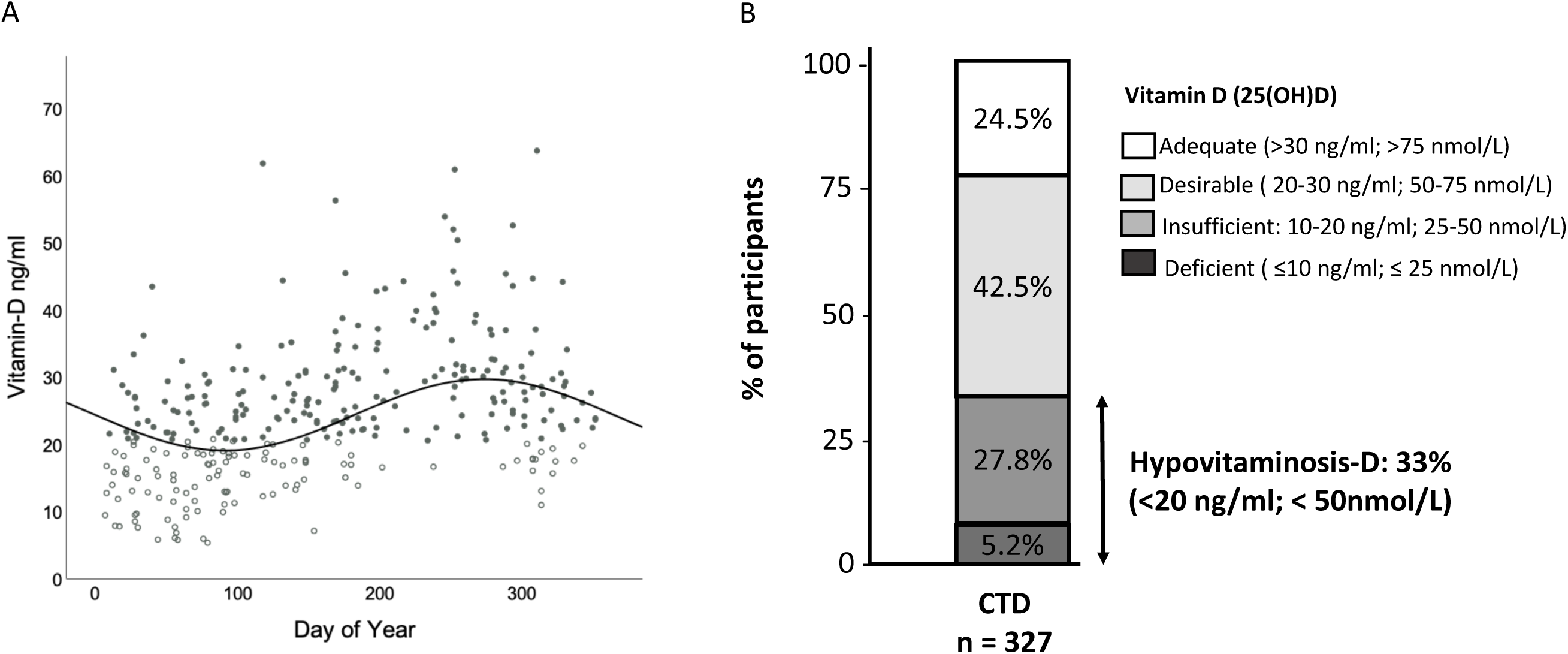
Levels of vitamin D (25(OH)D) in young individuals with chronic tic disorder (CTD). (A) Vitamin D serum measurements were plotted against 365 days of year (i.e. Jan 1^st^ = day 1). Reference line plotted is sine curve used to predict 25(OH)D levels. Predicted 25(OH)D levels peaks at day 240 (August 30^th^) and are lowest at day 58 (March 1^st^). Dots without fill are CTD patients with hypovitaminosis-D (<20 ng/ml; <50 nmol/L) and dots with black fill are CTD patients with 25(OH)D levels over 20 ng/ml. **(B) Clinical classification of serum vitamin D levels (25(OH)D) levels in young individuals with chronic tic disorder (CTD)**. 25(OH)D, 25-hydroxyvitamin D. Unit of measure 25(OH)D: nmol/L or ng/ml

One third of the participants (33.0%) presented with hypovitaminosis-D (25(OH)D <20 ng/ml; <50 nmol/L) and 5.2% were vitamin D deficient (<10 ng/ml; <25 nmol/L) (Figure 2B). The sinusoidal regression model used to adjust for seasonal variation was used to predict each participants’ serum 25(OH)D on any day of the year based on their residual value. This model predicted hypovitaminosis-D in 5.2% of the cohort year-round and 28.1% for 6 months of the year (December–May).

### No association between 25(OH)D levels and severity of tics

As shown in Table 2, using multilevel models, we found no significant association between tic severity and 25(OH)D levels (p = 0.86). This was also true for the YGTSS motor (p = 0.96) and vocal (p = 0.76). There was also no association between the Clinical Global Impression Scale Severity (CGI) of tics and 25(OH)D levels (p = 0.83).

**Table 2:** Mixed Model Analysis Severity Scores

| Independent Variable | Intercept | Fixed effects estimate for Vit D | SE | $p$ | 95% CI Lower | 95% CI Upper |
| --- | --- | --- | --- | --- | --- | --- |
| Tic Severity |  |  |  |  |  |  |
| YGTSS: Total | 13.11 | 0.01 | 0.05 | 0.86 | -0.09 | 0.11 |
| YGTSS Motor | 9.97 | 0.00 | 0.03 | 0.96 | -0.06 | 0.06 |
| YGTSS Vocal | 3.05 | 0.01 | 0.03 | 0.76 | -0.06 | 0.08 |
| CGI | 2.59 | 0.00 | 0.01 | 0.83 | -0.01 | 0.01 |
| OCD |  |  |  |  |  |  |
| CYBOCs Total | 4.13 | 0.05 | 0.05 | 0.34 | -0.05 | 0.16 |
| CYBOCs Obsessions | 0.00 | 0.02 | 0.03 | 0.38 | -0.03 | 0.08 |
| CYBOCs Compulsions | 4.36 | 0.03 | 0.03 | 0.41 | -0.04 | 0.10 |
| ADHD |  |  |  |  |  |  |
| **DSM-IV-TR Total** | **7.59** | **-0.10** | **0.03** | **0.00\*\*** | **-0.17** | **-0.04** |
| **DSM-IV-TR Inattentive** | **3.22** | **-0.06** | **0.02** | **0.00\*\*** | **-0.10** | **-0.02** |
| **DSM-IV-TR HI** | **4.42** | **-0.04** | **0.02** | **0.02\*** | **-0.07** | **-0.01** |
| **SNAP Total** | **17.23** | **-0.25** | **0.08** | **0.00\*\*** | **-0.42** | **-0.09** |
| **SNAP Inattentive** | **9.85** | **-0.17** | **0.05** | **0.00\*\*** | **-0.27** | **-0.08** |
| SNAP HI | 7.96 | -0.08 | 0.04 | 0.09 | -0.16 | 0.01 |

Abbreviations: YGTSS, Yale Global Tic Severity Scale; OCD, Obsessive-Compulsive Disorder; CY-BOCs, Children's Yale; ADHD, Attention deficit hyperactivity disorder; DSM IV, Diagnostic and statistical manual of mental disorders 4th ed., text revision; SNAP IV, Swanson, Nolan, and Pelham Questionnaire IV; HI, hyperactive-impulsive symptoms. The \*value represents the significant difference with $p < 0.05$ and \*\*value $p < 0.01$ .

### Association of 25(OH)D levels with comorbid ADHD but not OCD

OCD: As shown in Table 3, participants with lower 25(OH)D were less likely to have an OCD diagnosis (log odds = 0.04, s.e. = 0.03, 95% CI, 0.00 to 0.07; p = 0.04), results from the generalised linear mixed model predicts a 7% change in probability of an OCD diagnosis if a participants’ 25(OH)D level changes by one standard deviation either side of the mean for 25(OH)D (mean [SD] 25(OH)D, 25.21 ng/ml [8.59ng/ml]). However, we did not find an association between 25(OH)D levels and OCD symptom severity across the cohort, neither with the CY-BOCS total score (p = 0.34), nor the obsessions (p = 0.38) or compulsions (p = 0.41) subscales (Table 2).

**Table 3:** Generalised Linear Mixed Models: Vitamin D and Presence of Comorbidity

| Comorbidities | Intercept | Logit Odds VitD | SE | $p$ | 95% CI Lower | 95% CI Upper | Odds Ratio | Exp 95% CI Lower | Exp 95% CI Upper |
| --- | --- | --- | --- | --- | --- | --- | --- | --- | --- |
| OCD |  |  |  |  |  |  |  |  |  |
| **OCD/ no OCD** | **-1.83** | **0.04** | **0.02** | **0.04\*** | **0.00** | **0.07** | **1.04** | **1.00** | **1.07** |

ADHD
| ADHD/ no ADHD | 0.08 | -0.06 | 0.02 | 0.01** | -0.10 | -0.02 | 0.94 | 0.90 | 0.98 |
| --- | --- | --- | --- | --- | --- | --- | --- | --- | --- |

Generalised linear models were conducted to assess differences in serum 25-OHD between groups. The
\*\* value represents the significant difference with $p < 0.01$ .
The \*value represents the significant difference with $p < 0.05$ and \*\*value $p < 0.01$ .

#### ADHD

Participants with lower 25(OH)D were more likely to have an ADHD diagnosis (log odds = -0.06, s.e. = 0.02, 95% CI, -0.10 to -0.02; *p* < 0.01). These results estimate a 9% increase in the probability of an ADHD diagnosis if 25(OH)D levels are one standard deviation below the mean and a 7% decrease in probability for one standard deviation above the mean (mean [SD] vitamin D, 25.21 ng/ml [8.59ng/ml]). There was also a significant association between the parent-rated ADHD symptom severity based on the SNAP-IV (*β* = -0.25, s.e. = 0.08, *p* < 0.01), as well as for ADHD total symptom count based on the DSM-IV-TR and 25(OH)D levels (*β* = -0.10, s.e. = 0.03, *p* < 0.01) (Table 2) across the cohort. The association existed for the ADHD inattentive symptoms dimension, both for the SNAP-IV questionnaire (*β* = -0.17, s.e. = 0.05, *p* < 0.01) and the DSM-IV TR based counts (*β* = -0.06, s.e. = 0.02, *p* < 0.01). For hyperactive-impulsive symptoms, the DSM-IV-TR score was significant (*β* = -0.04, s.e. = 0.02, p = 0.02), whilst the SNAP score gave a non-significant trend (*β* = -0.08, s.e. = 0.04, p = 0.09) (Table 2).

## DISCUSSION

In this European cohort of children and adolescents with CTD we found hypovitaminosis-D (<20 ng/ml; <50 nmol/L) in 33.0% and severe 25(OH)D deficiency (<10 ng/ml; <25 nmol/L) in 5.2% of participants at the time of assessment. There was no association between 25(OH)D status and severity of tics or severity of OCD symptoms. However, a decrease in 25(OH)D, decreased the probability of an OCD diagnosis. Conversely, a decrease in 25(OH)D increased the probability of an ADHD diagnosis. In addition, lower 25(OH)D levels were significantly associated with the greater severity of ADHD symptoms in CTD patients. The inverse association was seen on both clinician and parent-rated measures of comorbid ADHD symptoms. Of note, an association was observed to a greater extent between serum 25(OH)D levels and inattentive symptoms than with hyperactive ADHD symptoms.

Our study highlighted the risk of hypovitaminosis-D and vitamin D deficiency in children and adolescents with CTD especially during winter months. Our multinational population was recruited from across Europe and Israel from 16 centres in nine countries. The analysis of vitamin D status included adequate adjustment for confounding factors including seasonality and site (which includes latitude) differences. In our study, 33.0% of participants presented with hypovitaminosis-D and 5.2% with vitamin D deficiency (<10ng/ml; 25 nmol/L) requiring treatment according to the National Institute of Health and Care Excellence (NICE) guidelines ^26^. Hypovitaminosis-D is common in Europe, however, prevalence rates vary significantly between age groups and limited data is available on children and adolescents ^27^. One limitation of this study was that there was no control group so we could not compare rates of hypovitaminosis-D in controls to participants with CTD. Nonetheless, raising awareness of the risk of vitamin D deficiency in children and adolescents with CTD is warranted as cost-effective prevention strategies exist such as altered diet, vitamin D supplements and changes in behaviour related to sun exposure.

Our cross-sectional study did not find an association between 25(OH)D status and tic severity in contrast to a recent study in Chinese children ^13^. This may be due to differences in ethnic background, genetic susceptibility and/or the type of tic disorder given that, in contrast to our study, 69% of the participants had a diagnosis of provisional tic disorder (hence, not chronic at the time of the study) and participants with psychiatric comorbidities were excluded.

A decrease in 25(OH)D by one standard deviation below the mean decreased the likelihood that a participant also had an OCD diagnosis by an estimate of 7%. However, the severity of comorbid OCD symptoms in children and adolescents with CTD was not linked to 25(OH)D status in our study. These incongruent findings do not allow for us to draw any clear conclusions on the relationship between OCD and 25(OH)D in this patient group. Few studies on vitamin D in OCD have so far been undertaken with inconclusive findings ^28^. However, two studies reported that OCD in patients with Paediatric Autoimmune Neuropsychiatric Disorder Associated with Streptococcal Infections (PANDAS) or Paediatric acute-onset neuropsychiatric syndrome (PANS) was associated with reduced vitamin D levels, which correlated with both number of streptococcal infections before PANDAS diagnosis and with infection recurrences ^29 30^.

Notably, we found an association between hypovitaminosis-D and the presence and severity of comorbid ADHD symptoms in children and adolescents with CTD. As we observed in our study, an estimate of *β* = -0.25 for SNAP-IV indicates that for an increase of 25(OH)D by one standard deviation above the mean (mean [SD] vitamin D, 25.21 ng/ml [8.59 ng/ml]), the score on SNAP-IV ADHD scale (range 0-54) decreased, on average, by 2.15. The physiological relevance of an increase in 8.59 ng/ml is not fully understood; current guidelines on 25(OH)D are based on its role in bone metabolism but less is known about the impact of changes in serum levels in relation to its involvement in immune mechanisms, brain development and neurotransmission.

ADHD has previously been linked to hypovitaminosis-D and to higher prevalence in regions with less sunshine ^31^. A recent meta-analysis, which analysed 10,334 children and adolescents, revealed that children with ADHD had lower levels of 25(OH)D than healthy children ^32^. In addition, a cross-sectional study reported that children with ADHD not only had lower levels of 25(OH)D but also lower serum levels of vitamin D receptor than controls ^33^.

In addition, we found that there was an association between 25(OH)D levels and inattentive symptoms to a greater extent than hyperactive ADHD symptoms in our cohort of children and adolescents with CTD. The findings were non-significant on SNAP-IV severity scale for hyperactive-impulsive symptoms and significant but with a smaller effect size than inattentive symptoms on the DSM-IV-TR symptom count. These findings are consistent with another recent cross-sectional study, which found no link between 25(OH)D levels and hyperactivity in children with ADHD, but a significant correlation between 25(OH)D levels and attention deficit as measured by the Conner’s questionnaire in ADHD ^34^. The neurobiological differences between ADHD subtypes are not well understood, although there is some suggestion of distinct alterations in underlying brain organization with greater hippocampal involvement in the inattentive ADHD-phenotype ^35^. One explanation for our findings could be that hypovitaminosis-D influences certain networks e.g. regions involved in attention and less so motor function, although more work is needed to validate this hypothesis.

The mechanism by which vitamin D status may be related to ADHD is currently unknown and warrants further study. There is evidence demonstrating the widespread presence of vitamin D receptors and 1a-hydroxylase (the enzyme responsible for the formation of the bioactive form of vitamin D) in the human brain, mainly in neuronal cells of the substantia nigra and hypothalamus but also hippocampus, prefrontal cortex and cingulated gyrus ^36^. Vitamin D is, therefore, thought to play an important role in brain ontogeny and mature brain function due to its neurotropic and neuroprotective effects and its impact on neurotransmission and neuroplasticity ^10 9^. There is an emerging literature indicating that vitamin D regulates dopamine neuron development and function ^37^. Supplementation with vitamin D3 was recently reported to increase serum dopamine levels in children with ADHD ^38^. Vitamin D may directly up-regulate the expression of tyrosine hydroxylase (a rate-limiting enzyme in dopamine synthesis) by binding to the nuclear vitamin D receptor and impact on the synthesis of serotonin in the brain ^39^. However, vitamin D also plays an important role in the regulation of inflammation, immunity, control of infection and the composition of the gut microbiome ^9^. These actions of vitamin D may also be of interest in the context of ADHD as there is support for the role of inflammation and autoimmunity in at least a subgroup of ADHD patients ^40^. Recent studies proposed a link to inflammatory and autoimmune diseases (e.g. asthma, atopy), neonatal infections, elevated serum interleukin levels (e.g. IL-6, IL-10), increased levels of anti-basal ganglia antibodies, antibodies against dopamine transporter and genes of the major histocompatibility complex ^40 41^.

Our cross-sectional study does not allow for drawing inferences about the direction of effects or causality between hypovitaminosis-D and comorbid ADHD symptoms in young people with CTD. Whether hypovitaminosis-D predisposes the development and severity of comorbid ADHD symptoms or vice versa (“reverse causation”) will need to be addressed. Prospective, longitudinal studies are therefore needed, which should also include information on diet, sunlight exposure, medication and screen for comorbid anxiety and depression. Vitamin D intervention studies may help assess whether vitamin D supplementation can help prevent ADHD or improve symptom severity in young individuals with CTD. There is evidence from recent randomised controlled trials to suggest that oral vitamin D supplementation in children with ADHD may improve ADHD symptoms ^42^.

In conclusion, our study is the first to investigate a relationship between 25(OH)D levels and severity of tics along with comorbid OCD and/or ADHD in young individuals with CTD. Our findings showed that 25(OH)D levels do not impact on tic severity or severity of OCD symptoms but are significantly associated with the presence of ADHD and severity of ADHD symptoms. Further research is needed on the role of hypovitaminosis-D on comorbid ADHD in this population as it may shed light on the underlying pathophysiology of ADHD and highlight novel strategies for intervention and prevention.

## Data Availability

The data can be obtained from the authors

## ACKNOWLEDGEMENTS

This project has received funding from the European Union’s Seventh Framework Programme for research, technological development and demonstration under grant agreement no 278367. The EMTICS collaborative group is added at the end of the manuscript.

The authors offer many thanks to the children and their parents who took part in this study. The authors want to thank Dr Sreeram Ramagopalan and Dr Giulio Disanto for help and discussion. This project is part of TransCampus, a joint partnership initiative of Technische Universitaet Dresden and King’s College London. This project has received funding from the European Union’s Seventh Framework Programme for research, technological development and demonstration under grant agreement no 278367.

We thank all members of the EMTICS collaborative group for their continued commitment to this project and in particular all colleagues at the various study centers who contributed to data collection and/or management: Julie E Bruun, Judy Grejsen, Christine L Ommundsen, Mette Rubæk (Capital Region Psychiatry, Copenhagen, Denmark); Stephanie Enghardt (TUD Dresden, Germany); Stefanie Bokemeyer, Christiane Driedger-Garbe, Cornelia Reichert (MHH Hannover, Germany); Jenny Schmalfeld (Lübeck University, Germany); Thomas Duffield (Munich, Germany); Franciska Gergye, Margit Kovacs, Reka Vidomusz (Vadaskert Budapest, Hungary); Miri Carmel, Silvana Fennig, Ella Gev, Nathan Keller, Elena Michaelovsky, Matan Nahon, Chen Regev, Tomer Simcha, Gill Smollan, Avi Weizman (Tel Aviv, Petah-Tikva, Israel); Giuseppe Gagliardi (Bari, Italy); Marco Tallon (Rome, Italy); Paolo Roazzi (Rome, Italy); Els van den Ban, Sebastian F.T.M. de Bruijn, Nicole Driessen, Andreas Lamerz, Marieke Messchendorp, Judith J.G. Rath, Nadine Schalk Deborah Sival, Noor Tromp, Frank Visscher and the Stichting Gilles de la Tourettes (UMCG Groningen, Netherlands); Maria Teresa Cáceres, Fátima Carrillo, Pilar Gómez-Garre, Laura Vargas (Seville, Spain); Maria Gariup (Barcelona Spain); Sara Stöber (Fürstenfeldbruck, Germany); and all who may not have been mentioned.

The EMTICS group members are Alan Apter^1^, Valentina Baglioni^2^, Juliane Ball^3^, Noa Benaroya-Milshtein^1^, Benjamin Bodmer^4^, Molly Bond^28^, Emese Bognar^5^, Bianka Burger^21^, Judith Buse^4^, Francesco Cardona^2^, Marta Correa Vela^6^, Andrea Dietrich^29^, Nanette M. Debes^7^, Maria Cristina Ferro^8^, Carolin Fremer^9^, Blanca Garcia-Delgar^10^, Mariangela Gulisano^8^, Annelieke Hagen^11,12^, Julie Hagstrøm^13^, Tammy J. Hedderly^14^, Isobel Heyman^15^, Pieter j Hoekstra ^29^, Chaim Huyser^11,12^, Marcos Madruga-Garrido^16^, Anna Marotta^17^, Davide Martino^18^, Ute-Christiane Meier^28,30^, Pablo Mir^6^, Natalie Moll^32^, Astrid Morer^10,19,20^, Norbert Mueller^33,34^, Kirsten Müller-Vahl^9^, Alexander Münchau^22^, Peter Nagy^5^, Valeria Neri^2^, Thaïra J.C. Openneer^23^, Alessandra Pellico^8^, Ángela Periañez Vasco^6^, Kerstin J. Plessen^13,24^, Cesare Porcelli^17^, Marina Redondo^10^, Renata Rizzo^8^, Veit Roessner^4^, Daphna Ruhrman^1^, Jaana M. L. Schnell ^21^, Anette Schrag^31^, Marcus J Schwarz^32^, Paola Rosaria Silvestri^2^, Liselotte Skov^26^, Tamar Steinberg^1^, Friederike Tagwerker Gloor^3^, Zsanett Tarnok^5^, Jennifer Tübing^27^, Victoria L. Turner^14^, Susanne Walitza^3^, Elif Weidinger ^21^, Martin L. Woods^14^

^1^Child and Adolescent Psychiatry Department, Schneider Children’s Medical Center of Israel, affiliated to Sackler Faculty of Medicine, Tel Aviv University, Petah-Tikva, Israel

^2^University La Sapienza of Rome, Department of Human Neurosciences, Rome, Italy

^3^Clinic of Child and Adolescent Psychiatry and Psychotherapy, University of Zurich, Zurich, Switzerland

^4^Department of Child and Adolescent Psychiatry, Faculty of Medicine of the TU Dresden, Dresden, Germany

^5^Vadaskert Child and Adolescent Psychiatric Hospital, Budapest, Hungary

^6^Unidad de Trastornos del Movimiento, Servicio de Neurología y Neurofisiología Clinica. Instituto de Biomedicina de Sevilla (IBiS), Hospital Universitario Virgen del Rocio/CSIC/Universidad de Sevilla, Seville, Spain

^7^Paediatric Department, Herlev University Hospital, Herlev, Denmark

^8^Child Neuropsychiatry Section, Department of Clinical and Experimental Medicine, School of Medicine, Catania University, Catania, Italy

^9^Clinic of Psychiatry, Socialpsychiatry and Psychotherapy, Hannover Medical School, Hannover, Germany

^10^Department of Child and Adolescent Psychiatry and Psychology, Institute of Neurosciences, Hospital Clinic Universitari, Barcelona, Spain

^11^De Bascule, Academic Center for Child and Adolescent Psychiatry, Amsterdam, The Netherlands

^12^Academic Medical Center, Department of Child and Adolescent Psychiatry, Amsterdam, The Netherlands

^13^Child and Adolescent Mental Health Center, Mental Health Services, Capital Region of Denmark and University of Copenhagen, Copenhagen, Denmark

^14^Evelina London Children’s Hospital GSTT, Kings Health Partners AHSC, London, UK

^15^Great Ormond Street Hospital for Children, and UCL Institute of Child Health, London, UK

^16^Sección de Neuropediatría, Instituto de Biomedicina de Sevilla (IBiS), Hospital Universitario Virgen del Rocío/CSIC/Universidad de Sevilla, Seville, Spain

^17^Azienda Sanitaria Locale di Bari, Mental Health Department, Child and Adolescent Service of Bari Metropolitan Area, Bari, Italy

^18^Department of Clinical Neurosciences, University of Calgary, Calgary, Canada

^19^Institut d’Investigacions Biomediques August Pi i Sunyer (IDIBAPS), Barcelona, Spain

^20^Centro de Investigacion en Red de Salud Mental (CIBERSAM), Instituto Carlos III, Spain

^21^ Department of Psychiatry and Psychotherapy, University Hospital, LMU Munich,

Munich, Germany

^22^Institute of Neurogenetics, University of Lübeck, Lübeck, Germany

^23^University of Groningen, University Medical Center Groningen, Department of Child and Adolescent Psychiatry, Groningen, The Netherlands

^24^Service of Child and Adolescent Psychiatry, Department of Psychiatry, University Hospital Lausanne, Lausanne, Switzerland

^25^Department of Clinical Neuroscience, UCL Institute of Neurology, University College London, London, UK

^26^concentris research management GmbH, Fürstenfeldbruck, Germany

^27^Department of Neurology, University of Lübeck, Lübeck, Germany

^28^Blizard Institute, Queen Mary University of London, Barts and The London School of Medicine and Dentistry, Department of Neuroscience and Trauma, Neuroinflammation and Immunopsychiatry Group, London, UK

^29^University of Groningen, University Medical Center Groningen, Department of Child and Adolescent Psychiatry, Groningen, The Netherlands

^30^Department of Psychological Medicine, Institute of Psychiatry, Psychology and Neuroscience, Kings College London, UK

^31^University College London, Institute of Neurology, London, UK

^32^Institute of Laboratory Medicine, University Hospital, LMU Munich, Germany

^33^ Department of Psychiatry and Psychotherapy, University Hospital, LMU Munich, Germany

^34^Marion von Tessin Memory-Zentrum, gGmbH, Munich, Germany

## DISCLOSURE

Dr Meier filed a patent “Biomarkers for inflammatory response”. All other authors reported no biomedical financial interest or potential conflict of interest.

## REFERENCES

1. American Psychiatric Association E. Diagnostic and statistical manual of mental disorders : DSM-5. 5th ed. Washington, D.C.: American Psychiatric Association 2013.

2. Leckman JF. Tourette’s syndrome. Lancet 2002;360(9345):1577–86. doi: 10.1016/S0140-6736(02)11526-1

3. Robertson MM. The prevalence and epidemiology of Gilles de la Tourette syndrome. Part 2: tentative explanations for differing prevalence figures in GTS, including the possible effects of psychopathology, aetiology, cultural differences, and differing phenotypes. J Psychosom Res 2008;65(5):473–86. doi: 10.1016/j.jpsychores.2008.03.007

4. van de Wetering BJ, Heutink P. The genetics of the Gilles de la Tourette syndrome: a review. J Lab Clin Med 1993;121(5):638–45.

5. Price RA, Kidd KK, Cohen DJ, et al. A twin study of Tourette syndrome. Arch Gen Psychiatry 1985;42(8):815–20.

6. Browne HA, Gair SL, Scharf JM, et al. Genetics of obsessive-compulsive disorder and related disorders. The Psychiatric clinics of North America 2014;37(3):319–35. doi: 10.1016/j.psc.2014.06.002

7. Larsson H, Chang Z, D’Onofrio BM, et al. The heritability of clinically diagnosed attention deficit hyperactivity disorder across the lifespan. Psychological medicine 2014;44(10):2223–9. doi: 10.1017/S0033291713002493

8. Eyles DW, Burne TH, McGrath JJ. Vitamin D, effects on brain development, adult brain function and the links between low levels of vitamin D and neuropsychiatric disease. Front Neuroendocrinol 2013;34(1):47–64. doi: 10.1016/j.yfrne.2012.07.001

9. Kocovska E, Gaughran F, Krivoy A, et al. Vitamin-D Deficiency As a Potential Environmental Risk Factor in Multiple Sclerosis, Schizophrenia, and Autism. Front Psychiatry 2017;8:47. doi: 10.3389/fpsyt.2017.00047

10. DeLuca GC, Kimball SM, Kolasinski J, et al. Review: the role of vitamin D in nervous system health and disease. Neuropathol Appl Neurobiol 2013;39(5):458–84. doi: 10.1111/nan.12020

11. Wacker M, Holick MF. Sunlight and Vitamin D: A global perspective for health. Dermatoendocrinol 2013;5(1):51–108. doi: 10.4161/derm.24494 [published Online First: 2014/02/05]

12. Ramagopalan SV, Dobson R, Meier UC, et al. Multiple sclerosis: risk factors, prodromes, and potential causal pathways. Lancet Neurol 2010;9(7):727–39. doi: 10.1016/S1474-4422(10)70094-6 [published Online First: 2010/07/09]

13. Li HH, Wang B, Shan L, et al. [Serum levels of 25-hydroxyvitamin D in children with tic disorders]. Zhongguo Dang Dai Er Ke Za Zhi 2017;19(11):1165-68. [published Online First: 2017/11/15]

14. Schrag A, Martino D, Apter A, et al. European Multicentre Tics in Children Studies (EMTICS): protocol for two cohort studies to assess risk factors for tic onset and exacerbation in children and adolescents. Eur Child Adolesc Psychiatry 2018 doi: 10.1007/s00787-018-1190-4 [published Online First: 2018/07/07]

15. Association AP. Diagnostic and statistical manual of mental disorders : DSM-IV-TR. 4th ed., text revision. ed. Washington, DC: American Psychiatric Association 2000.

16. Leckman JF, Riddle MA, Hardin MT, et al. The Yale Global Tic Severity Scale: initial testing of a clinician-rated scale of tic severity. J Am Acad Child Adolesc Psychiatry 1989;28(4):566–73. doi: 10.1097/00004583-198907000-00015

17. Guy W. Clinical Global Impression (CGI). ECDEU assessment manual for psychopharmacology.: U.S. Department of Health, Education, and Welfare, Rockville, MD, 1976.

18. Goodman WK, Price LH, Rasmussen SA, et al. The Yale-Brown Obsessive Compulsive Scale. II. Validity. Arch Gen Psychiatry 1989;46(11):1012–6.

19. Swanson JM, Kraemer HC, Hinshaw SP, et al. Clinical relevance of the primary findings of the MTA: success rates based on severity of ADHD and ODD symptoms at the end of treatment. J Am Acad Child Adolesc Psychiatry 2001;40(2):168–79. doi: 10.1097/00004583-200102000-00011

20. Holick MF, Binkley NC, Bischoff-Ferrari HA, et al. Evaluation, treatment, and prevention of vitamin D deficiency: an Endocrine Society clinical practice guideline. The Journal of clinical endocrinology and metabolism 2011;96(7):1911–30. doi: 10.1210/jc.2011-0385

21. Ross AC, Manson JE, Abrams SA, et al. The 2011 report on dietary reference intakes for calcium and vitamin D from the Institute of Medicine: what clinicians need to know. The Journal of clinical endocrinology and metabolism 2011;96(1):53–8. doi: 10.1210/jc.2010-2704

22. Mowry EM, Krupp LB, Milazzo M, et al. Vitamin D status is associated with relapse rate in pediatric-onset multiple sclerosis. Ann Neurol 2010;67(5):618–24. doi: 10.1002/ana.21972

23. Raudenbush SW, Bryk AS. Hierarchical linear models : applications and data analysis methods. 2nd ed. ed. Thousand Oaks, CA ; London: Sage Publications 2002.

24. Enders CK, Tofighi D. Centering predictor variables in cross-sectional multilevel models: a new look at an old issue. Psychol Methods 2007;12(2):121–38. doi: 10.1037/1082-989X.12.2.121

25. Holick MF. Vitamin D status: measurement, interpretation, and clinical application. Ann Epidemiol 2009;19(2):73–8. doi: 10.1016/j.annepidem.2007.12.001

26. NICE. Vitamin D deficiency in children: National Institute for Health and Care Excellence; 2018 [Available from: https://cks.nice.org.uk/vitamin-d-deficiency-in-children#!scenario.

27. Braegger C, Campoy C, Colomb V, et al. Vitamin D in the healthy European paediatric population. Journal of pediatric gastroenterology and nutrition 2013;56(6):692–701. doi: 10.1097/MPG.0b013e31828f3c05

28. Yazici KU, Percinel Yazici I, Ustundag B. Vitamin D levels in children and adolescents with obsessive compulsive disorder. Nord J Psychiatry 2018;72(3):173–78. doi: 10.1080/08039488.2017.1406985 [published Online First: 2017/11/23]

29. Celik G, Tas D, Tahiroglu A, et al. Vitamin D Deficiency in Obsessive-Compulsive Disorder Patients with Pediatric Autoimmune Neuropsychiatric Disorders Associated with Streptococcal Infections: A Case Control Study. Noro Psikiyatr Ars 2016;53(1):33–37. doi: 10.5152/npa.2015.8763 [published Online First: 2017/04/01]

30. Stagi S, Lepri G, Rigante D, et al. Cross-Sectional Evaluation of Plasma Vitamin D Levels in a Large Cohort of Italian Patients with Pediatric Autoimmune Neuropsychiatric Disorders Associated with Streptococcal Infections. J Child Adolesc Psychopharmacol 2018;28(2):124–29. doi: 10.1089/cap.2016.0159 [published Online First: 2017/11/08]

31. Arns M, van der Heijden KB, Arnold LE, et al. Geographic variation in the prevalence of attention-deficit/hyperactivity disorder: the sunny perspective. Biol Psychiatry 2013;74(8):585–90. doi: 10.1016/j.biopsych.2013.02.010 [published Online First: 2013/03/26]

32. Khoshbakht Y, Bidaki R, Salehi-Abargouei A. Vitamin D Status and Attention Deficit Hyperactivity Disorder: A Systematic Review and Meta-Analysis of Observational Studies. Adv Nutr 2018;9(1):9–20. doi: 10.1093/advances/nmx002 [published Online First: 2018/02/14]

33. Sahin N, Altun H, Kurutas EB, et al. Vitamin D and vitamin D receptor levels in children with attention-deficit/hyperactivity disorder. Neuropsychiatr Dis Treat 2018;14:581–85. doi: 10.2147/NDT.S158228 [published Online First: 2018/03/03]

34. Reinehr T, Langrock C, Hamelmann E, et al. 25-Hydroxvitamin D concentrations are not lower in children with bronchial asthma, atopic dermatitis, obesity, or attention-deficient/hyperactivity disorder than in healthy children. Nutr Res 2018;52:39–47. doi: 10.1016/j.nutres.2018.01.002 [published Online First: 2018/05/17]

35. Saad JF, Griffiths KR, Kohn MR, et al. Regional brain network organization distinguishes the combined and inattentive subtypes of Attention Deficit Hyperactivity Disorder. Neuroimage Clin 2017;15:383–90. doi: 10.1016/j.nicl.2017.05.016 [published Online First: 2017/06/06]

36. Eyles DW, Smith S, Kinobe R, et al. Distribution of the vitamin D receptor and 1 alpha-hydroxylase in human brain. J Chem Neuroanat 2005;29(1):21–30. doi: 10.1016/j.jchemneu.2004.08.006 [published Online First: 2004/12/14]

37. Pertile RAN, Cui X, Hammond L, et al. Vitamin D regulation of GDNF/Ret signaling in dopaminergic neurons. FASEB J 2018;32(2):819–28. doi: 10.1096/fj.201700713R [published Online First: 2017/10/12]

38. Seyedi M, Gholami F, Samadi M, et al. The Effect of Vitamin D3 Supplementation on Serum BDNF, Dopamine and Serotonin in Children with Attention-Deficit/Hyperactivity Disorder. CNS Neurol Disord Drug Targets 2019 doi: 10.2174/1871527318666190703103709 [published Online First: 2019/07/05]

39. Patrick RP, Ames BN. Vitamin D and the omega-3 fatty acids control serotonin synthesis and action, part 2: relevance for ADHD, bipolar disorder, schizophrenia, and impulsive behavior. FASEB J 2015;29(6):2207–22. doi: 10.1096/fj.14-268342 [published Online First: 2015/02/26]

40. Leffa DT, Torres ILS, Rohde LA. A Review on the Role of Inflammation in Attention-Deficit/Hyperactivity Disorder. Neuroimmunomodulation 2018:1–6. doi: 10.1159/000489635 [published Online First: 2018/06/06]

41. Odell JD, Warren RP, Warren WL, et al. Association of genes within the major histocompatibility complex with attention deficit hyperactivity disorder. Neuropsychobiology 1997;35(4):181–6. doi: 10.1159/000119342

42. Gan J, Galer P, Ma D, et al. The Effect of Vitamin D Supplementation on Attention-Deficit/Hyperactivity Disorder: A Systematic Review and Meta-Analysis of Randomized Controlled Trials. J Child Adolesc Psychopharmacol 2019 doi: 10.1089/cap.2019.0059 [published Online First: 2019/08/01]

